# Emergence and rapid spread of a new severe acute respiratory syndrome-related coronavirus 2 (SARS-CoV-2) lineage with multiple spike mutations in South Africa

**DOI:** 10.1101/2020.12.21.20248640

**Authors:** Houriiyah Tegally, Eduan Wilkinson, Marta Giovanetti, Arash Iranzadeh, Vagner Fonseca, Jennifer Giandhari, Deelan Doolabh, Sureshnee Pillay, Emmanuel James San, Nokukhanya Msomi, Koleka Mlisana, Anne von Gottberg, Sibongile Walaza, Mushal Allam, Arshad Ismail, Thabo Mohale, Allison J Glass, Susan Engelbrecht, Gert Van Zyl, Wolfgang Preiser, Francesco Petruccione, Alex Sigal, Diana Hardie, Gert Marais, Marvin Hsiao, Stephen Korsman, Mary-Ann Davies, Lynn Tyers, Innocent Mudau, Denis York, Caroline Maslo, Dominique Goedhals, Shareef Abrahams, Oluwakemi Laguda-Akingba, Arghavan Alisoltani-Dehkordi, Adam Godzik, Constantinos Kurt Wibmer, Bryan Trevor Sewell, José Lourenço, Luiz Carlos Junior Alcantara, Sergei L Kosakovsky Pond, Steven Weaver, Darren Martin, Richard J Lessells, Jinal N Bhiman, Carolyn Williamson, Tulio de Oliveira

**Affiliations:** KwaZulu-Natal Research Innovation and Sequencing Platform (KRISP), Department of Laboratory Medicine & Medical Sciences, University of KwaZulu-Natal, Durban, South Africa; Laboratório de Genética Celular e Molecular, Universidade Federal de Minas Gerais, Belo Horizonte, Minas Gerais, Brazil; Laboratorio de Flavivirus, Fundacao Oswaldo Cruz, Rio de Janeiro, Brazil; Computational Biology Division, Department of Integrative Biomedical Sciences, University of Cape Town, Cape Town, 7925, South Africa; Division of Medical Virology, Institute of Infectious Disease and Molecular Medicine, University of Cape Town, Cape Town, South Africa; Discipline of Virology, University of KwaZulu-Natal, School of Laboratory Medicine and Medical Sciences and National Health Laboratory Service, Durban, South Africa; National Health Laboratory Service, Johannesburg, South Africa; Centre for the AIDS Programme of Research in South Africa (CAPRISA), Durban, South Africa; National Institute for Communicable Diseases of the National Health Laboratory Service, Johannesburg, South Africa; School of Pathology, Faculty of Health Sciences, University of the Witwatersrand, Johannesburg, South Africa; School of Public Health, Faculty of Health Sciences, University of the Witwatersrand, Johannesburg, South Africa; Department of Molecular Pathology, Lancet Laboratories, Johannesburg, South Africa; Division of Medical Virology at NHLS Tygerberg Hospital and Faculty of Medicine and Health Sciences, Stellenbosch University, Cape Town, South Africa; Centre for Quantum Technology, University of KwaZulu-Natal, Durban, South Africa; National Institute for Theoretical Physics (NITheP), KwaZulu-Natal, South Africa; Africa Health Research Institute, Durban, South Africa; School of Laboratory Medicine and Medical Sciences, University of KwaZulu-Natal, Durban, South Africa; Max Planck Institute for Infection Biology, Berlin, Germany; Division of Medical Virology at NHLS Groote Schuur Hospital, University of Cape Town, Cape Town, South Africa; Centre for Infectious Disease Epidemiology and Research, University of Cape Town, Cape Town, South Africa; Western Cape Government: Health, Cape Town, South Africa; Molecular Diagnostics Services, Durban, South Africa; Department of Quality Leadership, Netcare Hospitals, Johannesburg, South Africa; Division of Virology at NHLS Universitas Academic Laboratories, University of The Free State, Bloemfontein, South Africa; National Health Laboratory Service, Port Elizabeth, South Africa; Department of Laboratory Medicine and Pathology, Faculty of Health Sciences, Walter Sisulu University, Mthatha, South Africa; Division of Medical Virology, Department of Pathology, University of Cape Town, Cape Town, South Africa; Division of Biomedical Sciences, University of California Riverside School of Medicine, Riverside, California, USA; Structural Biology Research Unit, Department of Integrative Biomedical Sciences, University of Cape Town, Rondebosch, South Africa; Department of Zoology, University of Oxford, Oxford, United Kingdom; Institute for Genomics and Evolutionary Medicine, Temple University, Philadelphia, Pennsylvania, USA; Department of Global Health, University of Washington, Seattle, USA

## Abstract

Continued uncontrolled transmission of the severe acute respiratory syndrome-related coronavirus 2 (SARS-CoV-2) in many parts of the world is creating the conditions for significant virus evolution. Here, we describe a new SARS-CoV-2 lineage (501Y.V2) characterised by eight lineage-defining mutations in the spike protein, including three at important residues in the receptor-binding domain (K417N, E484K and N501Y) that may have functional significance. This lineage emerged in South Africa after the first epidemic wave in a severely affected metropolitan area, Nelson Mandela Bay, located on the coast of the Eastern Cape Province. This lineage spread rapidly, becoming within weeks the dominant lineage in the Eastern Cape and Western Cape Provinces. Whilst the full significance of the mutations is yet to be determined, the genomic data, showing the rapid displacement of other lineages, suggest that this lineage may be associated with increased transmissibility.

## Introduction

Severe acute respiratory syndrome-related coronavirus 2 (SARS-CoV-2) emerged in 2019 and spread rapidly around the world, causing over 70 million recorded cases of coronavirus disease (COVID-19) and over 1.6 million deaths by mid-December 2020. The failure of public health measures to contain the spread of the virus within and between countries has given rise to a large number of virus lineages across the world. Open genomic surveillance data sharing and collaborative online platforms have enabled real-time tracking of the emergence and spread of these lineages^1,2^.

To date there has been relatively limited evidence of SARS-CoV-2 mutations that have had a significant functional effect on the virus. One mutation in the spike protein (D614G) emerged early in the epidemic and spread rapidly through Europe and North America in particular Several lines of evidence now suggest that SARS-CoV-2 variants carrying this mutation have increased transmissibility^3-6^. Later in the epidemic, lineages with a N439K mutation in the spike receptor-binding domain (RBD) emerged independently in different European countries and in the United States. This mutation is associated with escape from monoclonal antibody (mAb) and polyclonal serum mediated neutralization^7^.

South Africa has been the most severely affected country in Africa, with over 56 000 excess natural deaths having occurred by 8 December (approximately 950 per million population)^8^. We have previously described the introduction and spread of several SARS-CoV-2 lineages, and the emergence of unique South African lineages during the early phase of the epidemic^9,10^. Here, we now describe the emergence and spread of a new SARS-CoV-2 lineage harbouring multiple nonsynonymous spike mutations, including mutations at key sites in the RBD (K417N, E484K and N501Y) that may have functional significance. We demonstrate that this lineage emerged after the first epidemic wave in the worst affected metropolitan area within the Eastern Cape (EC) Province. This was followed by rapid spread of this lineage to the extent that it has now become the dominant lineage in EC and Western Cape (WC) Provinces.

## Methods

### Epidemiological dynamics

We analysed daily cases of SARS-CoV-2 in South Africa up to 14 December 2020 from publicly released data provided by the National Department of Health (NDoH) and the National Institute for Communicable Diseases. This was accessible through the repository of the Data Science for Social Impact Research Group at the University of Pretoria (https://github.com/dsfsi/covid19za)^11,12^. The NDoH releases daily updates on the number of confirmed new cases, deaths and recoveries, with a breakdown by province. We also mapped excess deaths in each province and in South Africa as a whole on to general epidemiological data to determine the extent of potential under-reporting of case numbers and gauge the severity of the epidemic. Excess deaths here are defined as the excess natural deaths (in individuals aged 1 year and above) relative to the value predicted from 2018 and 2019 data, setting any negative excesses to zero. We obtained the data from the Report on Weekly Deaths from the South Africa Medical Research Council Burden of Disease Research Unit^8^. We generated estimates for the effective reproduction number (Re) of SARS-CoV-2 in South Africa from the covid-19-re data repository (https://github.com/covid-19-Re/dailyRe-Data) as of 14 December 2020^13^.

### Sampling of SARS-CoV-2

As part of the Network for Genomic Surveillance in South Africa (NGS-SA)^14^, five sequencing hubs receive randomly selected samples for sequencing every week according to approved protocols at each site. These samples include remnant nucleic acid extracts or remnant nasopharyngeal and oropharyngeal swab samples from routine diagnostic SARS-CoV-2 PCR testing from public and private laboratories in South Africa. In response to a rapid resurgence of COVID-19 in EC and the Garden Route District of WC in November, we enriched our routine sampling with additional samples from those areas. In total, we received samples from over 50 health facilities in the EC and WC (Suppl Table 1, Suppl Fig. S1, Suppl Fig. S2).

### Ethical Statement

The project was approved by University of KwaZulu-Natal Biomedical Research Ethics Committee. Protocol reference number: BREC/00001510/2020. Project title: Spatial and genomic monitoring of COVID-19 cases in South Africa. This project was also approved by University of the Witwatersrand Human Research Ethics Committee. Clearance certificate number: M180832. Project title: Surveillance for outpatient influenza-like illness and asymptomatic virus colonization in South Africa. Sequence data from the Western Cape was approved by the Stellenbosch University HREC Reference No: N20/04/008_COVID-19.

Project Title: COVID-19: sequencing the virus from South African patients.

### Whole genome sequencing and genome assembly

cDNA synthesis was performed on the extracted RNA using random primers followed by gene specific multiplex PCR using the ARTIC V3 protocol^15^. Briefly, extracted RNA was converted to cDNA using the Superscript IV First Strand synthesis system (Life Technologies, Carlsbad, CA) and random hexamer primers. SARS-CoV-2 whole genome amplification was performed by multiplex PCR using primers designed on Primal Scheme (http://primal.zibraproject.org/) to generate 400bp amplicons with an overlap of 70bp that covers the 30Kb SARS-CoV-2 genome. PCR products were cleaned up using AmpureXP purification beads (Beckman Coulter, High Wycombe, UK) and quantified using the Qubit dsDNA High Sensitivity assay on the Qubit 4.0 instrument (Life Technologies Carlsbad, CA).

We then used the Illumina® Nextera Flex DNA Library Prep kit according to the manufacturer’s protocol to prepare indexed paired end libraries of genomic DNA. Sequencing libraries were normalized to 4nM, pooled and denatured with 0.2N sodium acetate. 12pM sample library was spiked with 1% PhiX (PhiX Control v3 adapter-ligated library used as a control). We sequenced libraries on a 500-cycle v2 MiSeq Reagent Kit on the Illumina MiSeq instrument (Illumina, San Diego, CA, USA). We have previously published full details of the amplification and sequencing protocol^16,17^.

We assembled paired-end fastq reads using Genome Detective 1.126 (https://www.genomedetective.com) and the Coronavirus Typing Tool^18^. We polished the initial assembly obtained from Genome Detective by aligning mapped reads to the references and filtering out low-quality mutations using bcftools 1.7-2 mpileup method. Mutations were confirmed visually with bam files using Geneious software (Biomatters Ltd, New Zealand). All of the sequences were deposited in GISAID (https://www.gisaid.org/), and the GISAID accession IDs are included as part of Suppl Table 1. Raw reads for our sequences have also been deposited at the National Center for Biotechnology Information Sequence Read Archive.

### Quality control of South African genomic sequences

We retrieved all South African SARS-CoV-2 genotypes from the GISAID database as of 14 December 2020 (N=2704). Prior to phylogenetic reconstruction, we removed low quality sequences from this dataset. We filtered out genotypes that did not pass standard quality assessment parameters employed in NextClade (https://clades.nextstrain.org). We filtered out 99 South African genotypes due to low coverage, and a further 16 due to poor sequence quality. We therefore analyzed a total of 2589 South African genotypes. We also retrieved a global reference dataset (N=2592). This was selected from the Nextstrain global reference dataset, plus the five most similar sequences to each of the South African sequences as defined by a local BLAST search.

### Phylogenetic analysis

We initially analyzed South African genotypes against the global reference dataset using a custom pipeline based on a local version of NextStrain^2^. The pipeline contains several python scripts that manage the analysis workflow. It performs alignment of genotypes in MAFFT^19^, phylogenetic tree inference in IQ-Tree^20^, tree dating and ancestral state construction and annotation (https://github.com/nextstrain/ncov).

The initial phylogenetic analysis allowed us to identify a large cluster of sequences (n=190) with multiple spike mutations. We extracted this cluster and constructed a preliminary maximum likelihood (ML) tree in IQ-tree, together with eight basal sequences from the region sampled June-September 2020. We inspected this ML tree in TempEst v1.5.3 for the presence of a temporal (i.e. molecular clock) signal. Linear regression of root-to-tip genetic distances against sampling dates indicated that SARS-CoV-2 sequences evolved in a relatively strong clock-like manner (correlation coefficient=0.65, R^2^=0.41) (Suppl Fig. S2).

We then estimated time-calibrated phylogenies using the Bayesian software package BEASTv.1.10.4. For this analysis, we employed the strict molecular clock model, the HKY+I, nucleotide substitution model and the exponential growth coalescent model^21^. We computed Markov chain Monte Carlo (MCMC) in duplicate runs of 100 million states each, sampling every 10 000 steps. Convergence of MCMC chains was checked using Tracer v.1.7.1^22^. Maximum clade credibility trees were summarised from the MCMC samples using TreeAnnotator after discarding 10% as burn-in.

### Phylogeographic analysis

To model phylogenetic diffusion of the new cluster across the country, we used a flexible relaxed random walk (RRW) diffusion model that accommodates branch-specific variation in rates of dispersal with a Cauchy distribution^23^. For each sequence, latitude and longitude were attributed to the health facility at which the diagnostic sample was obtained, or, if that information was not available, to a point randomly sampled within the local area or district of origin.

As above, MCMC chains were run in duplicate for 100 million generations and sampled every 10 000 steps, with convergence assessed using Tracer v1.7.1. Maximum clade credibility trees were summarized using TreeAnnotator after discarding 10% as burn-in. We used the R package “seraphim” to extract and map spatiotemporal information embedded in posterior trees.

### Lineage classification

We used the dynamic lineage classification method proposed by Rambault et al. via the Phylogenetic Assignment of named Global Outbreak LINeages (PANGOLIN) software suite (https://github.com/hCoV-2019/pangolin)^24^. This is aimed at identifying the most epidemiologically important lineages of SARS-CoV-2 at the time of analysis, allowing researchers to monitor the epidemic in a particular geographical region. For the new cluster identified in South Africa in this study, we have assigned it the name 501Y.V2 until PANGOLIN reflects the lineage in a future update.

### Selection analysis

To identify which, if any, of the observed mutations in the spike protein was most likely to increase viral fitness, we used the natural selection analysis of SARS-CoV-2 pipeline (https://observablehq.com/@spond/revised-sars-cov-2-analytics-page). This pipeline examines the entire global SARS-CoV-2 nucleotide sequence dataset for evidence of: (i) polymorphisms having arisen in multiple epidemiologically unlinked lineages that have statistical support for non-neutral evolution (Mixed Effects Model of Evolution, MEME)^25^, (ii) sites where these polymorphisms have support for a greater than expected ratio of non-synonymous:synonymous nucleotide substitution rates on internal branches of the phylogenetic tree (Fixed Effects Likelihood, FEL)^26^, and (iii) whether these polymorphisms have increased in frequency in the regions of the world where they have occurred.

### Structural modelling

We modelled the spike protein based on the Protein Data Bank coordinate set 7a94, showing the first step of the S protein trimer activation with one RBD domain in the up position, bound to the hACE2 receptor^27^. We used the Pymol program (The PyMOL Molecular Graphics System, Version 2.2.0, Schrödinger, LLC.) for visualization.

## Results

### Epidemic dynamics in South Africa

The second SARS-CoV-2 epidemic wave in South Africa began around October 2020, just weeks after a trough in daily recorded cases following the first peak (Fig. 1A)^28^. The country’s estimated effective reproduction number, Re, increased to above 1 at the end of October, indicating a growing epidemic, coinciding with a steady rise in daily cases. At the peak of the national epidemic in mid-July there were over 13 000 confirmed cases per day and almost 7000 excess deaths per week. The epidemiological profile in the three provinces that are the focus of this analysis (EC, WC and KZN) were broadly similar, although WC had an earlier and flatter peak in the first wave (Fig. 1B-D). At the end of the first wave in early September, there had been over 10 000 excess deaths in EC (1510 per million population), the highest for any province (Suppl Fig. S3). Although there was a plateau in cases following the first wave, this was noticeably short in EC and by early October there was a second phase of exponential growth, associated with an increase in deaths at a similar rate to the first wave (Fig. 1B). PCR test positivity rate data at a local municipality level shows very high levels (>20%) in Nelson Mandela Bay from mid-October and then rapidly rising levels in surrounding areas through October and November (Suppl Fig. S4). The resurgence of the daily case counts at an exponential rate happened later for WC and KZN than for EC (Fig. 1C-D). By early December, all three provinces were experiencing a second wave, and new cases in WC had already surpassed the peak of the first wave.

**Figure 1.**
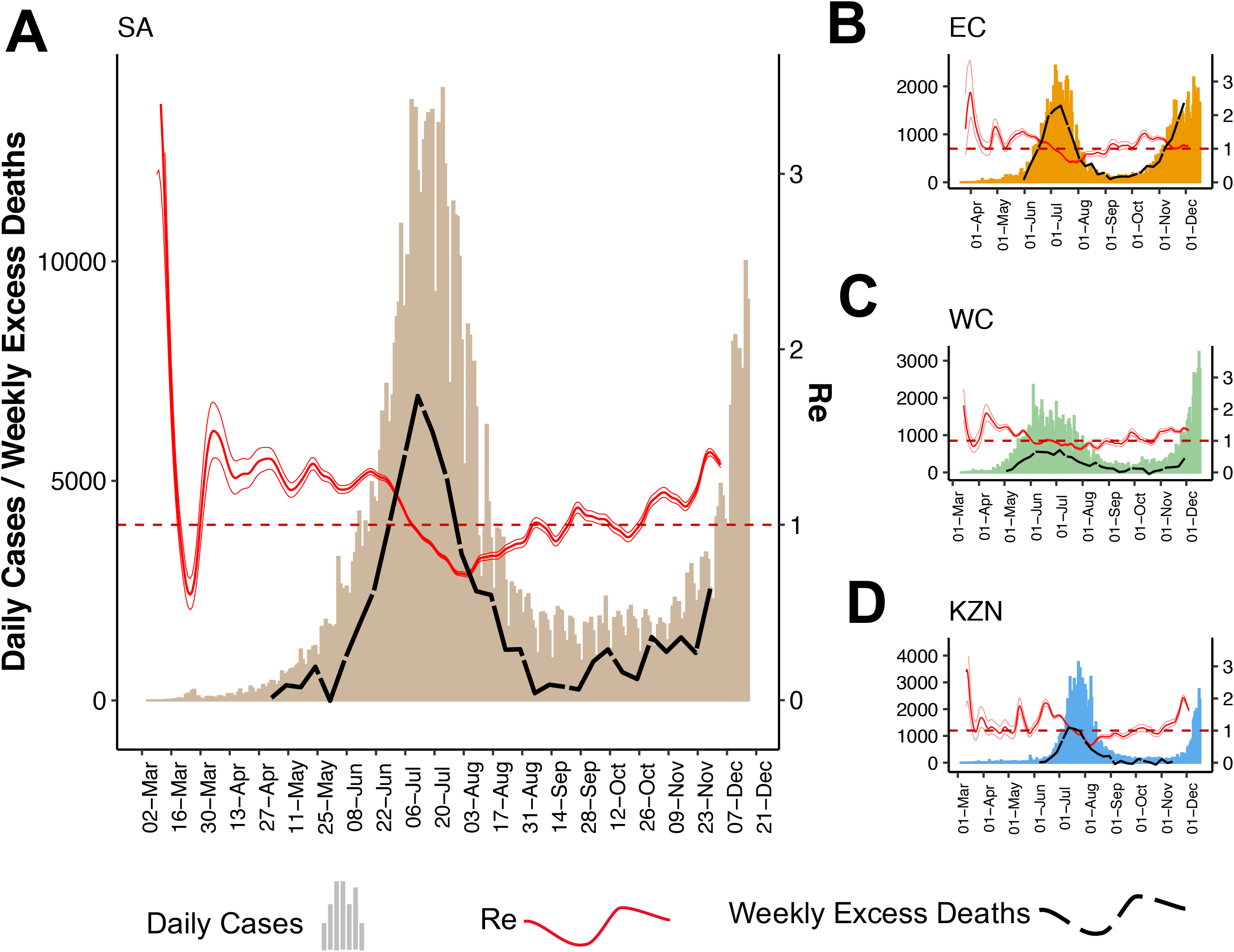
SARS-CoV-2 epidemiological dynamics in South-Africa (A), and the three provinces under study, Eastern Cape (B), Western Cape (C), and KwaZulu-Natal (D). The histograms show the number of daily confirmed COVID-19 cases in each region (mapped to left y-axis). Fluctuations to daily Re estimates are shown in red (mapped to right y-axis), with a cut-off for R=1 shown as the red broken line. Weekly excess deaths in each region are shown with the black broken lines (mapped to the left y-axis).

### Phylogenetic and phylogeographic analysis

The early and rapid resurgence of the epidemic in parts of the EC and WC prompted intensification of genomic surveillance by the NGS-SA, including sampling in and around Nelson Mandela Bay in EC, and in the neighbouring Garden Route district of WC (Suppl Fig. S5.). We analysed 2589 SARS-CoV-2 whole genomes from South Africa collected between 5 March and 25 November 2020. We estimated preliminary maximum likelihood (ML) and molecular clock phylogenies for a dataset containing as many global reference genomes (a subset of the tree is shown in Fig. 2A). We identified a new monophyletic cluster (501Y.V2) containing 190 sequences from samples collected between 15 October and 25 November in KZN, EC and WC (Fig. 2B). By early November, the 501Y.V2 lineage had superseded the three main South African lineages (B.1.1.54, B.1.1.56 and C.1) that were circulating during the first epidemic wave, and rapidly became the dominant lineage in samples from EC and WC (Fig. 2C, Suppl Fig. S6,S7).

**Figure 2.**
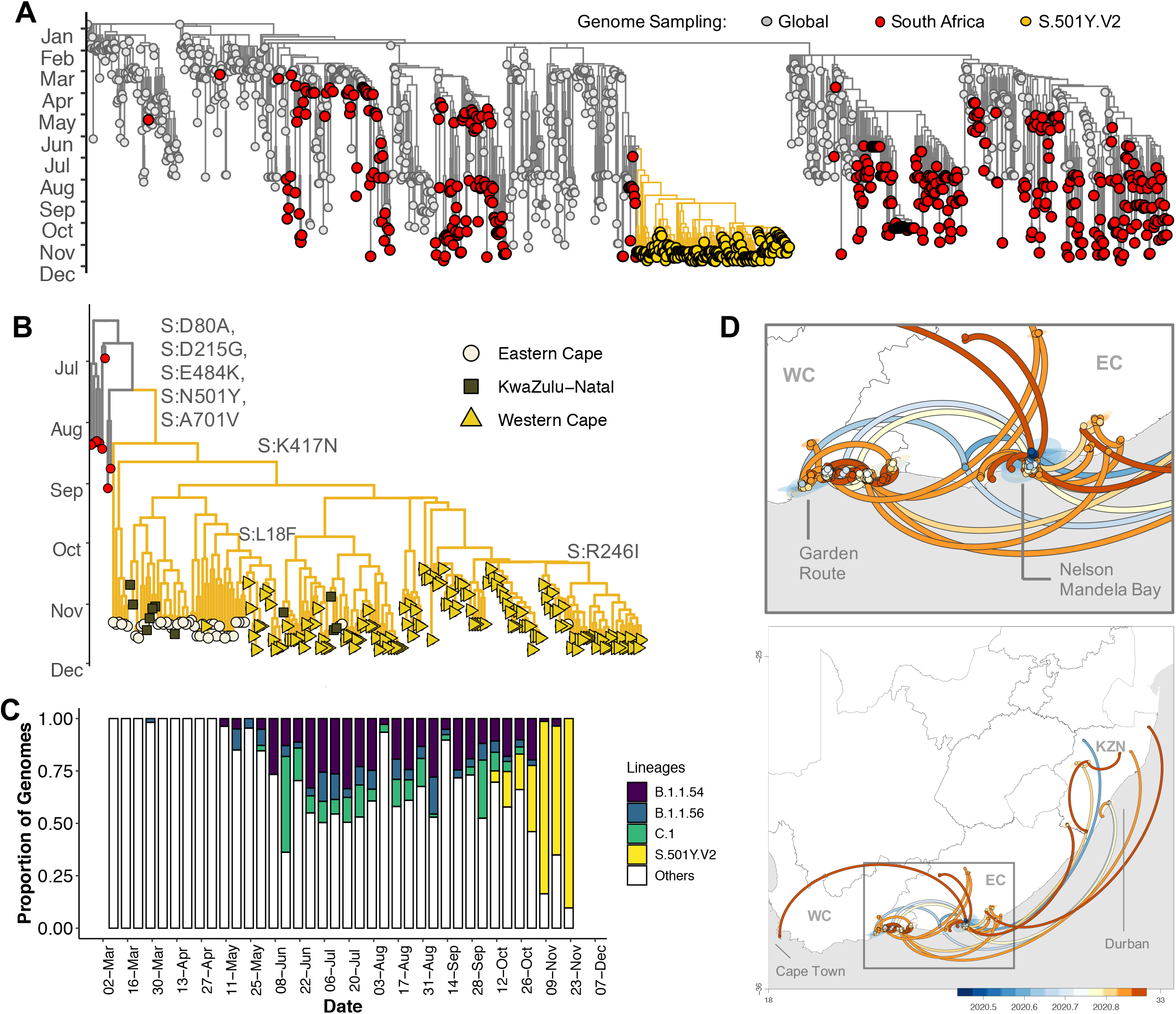
Evolution and spread of the S.501Y.V2 cluster in South Africa. A) Time-resolved maximum clade credibility phylogeny of 1350 SARS-CoV-2 sequences, 647 of which are from South Africa (red). The new SARS-CoV-2 cluster is highlighted in yellow. B) Time-resolved maximum clade credibility phylogeny of S.501Y.V2 cluster, with province location indicated. Mutations characterizing the cluster are highlighted at each branch where they first emerged. C) Frequency and distribution of SARS-CoV-2 lineages circulating in South Africa over time. D) Spatiotemporal reconstruction of the spread of the S.501Y.V2 cluster in South Africa during the second epidemic wave. Circles represent nodes of the maximum clade credibility phylogeny and are colored according to their inferred time of occurrence. Shaded areas represent the 80% highest posterior density interval and depict the uncertainty of the phylogeographic estimates for each node. Solid curved lines denote the links between nodes and the directionality of movement. The date scale goes from July 2020 (2020.5) to November 2020 (2020.9)

Spatiotemporal phylogeographic analysis suggests that the 501Y.V2 lineage emerged in early August (early July – end August 2020, 95% highest posterior density) in Nelson Mandela Bay. Initial spread to the Garden Route District of WC was then followed by more diffuse spread from both of those areas to other regions of EC, and more recently to the City of Cape Town and several locations in KZN (Fig. 2D).

### Mutational profile

At the point of first sampling on 15 October this lineage had, in addition to D614G, five other non-synonymous mutations in the spike protein, namely D80A, D215G, E484K, N501Y and A701V (Fig. 2B, Fig. 3A, Suppl Fig. S8). Three further spike mutations emerged by the end of November: L18F, R246I and K417N. We also observe a potential deletion of three amino acids at L242_244L, seen in samples extracted and generated in various laboratories across the network, however this site is still unresolved as it is disputed with a L242H mutation in our sequences. Three of the spike mutations are at key residues in the RBD (N501Y, E484K and K417N), three are in the N-terminal domain (L18F, D80A and D215G) and one is in loop 2 (A701V) (Suppl Fig. S9). While the variants appeared in a varying proportion of the sampled genomes and showed changing frequency levels with time, the RBD mutations seem to become fixed in our sampling set, present in almost all the samples, and consistently high in frequency across time (Fig. 3A-B). Compared to the three largest lineages circulating in SA previously, S501.V2 shows marked hypermutation both in the whole genomes and the spike regions, including nonsynonymous variants leading to amino acid changes (Fig. 3C).

**Figure 3.**
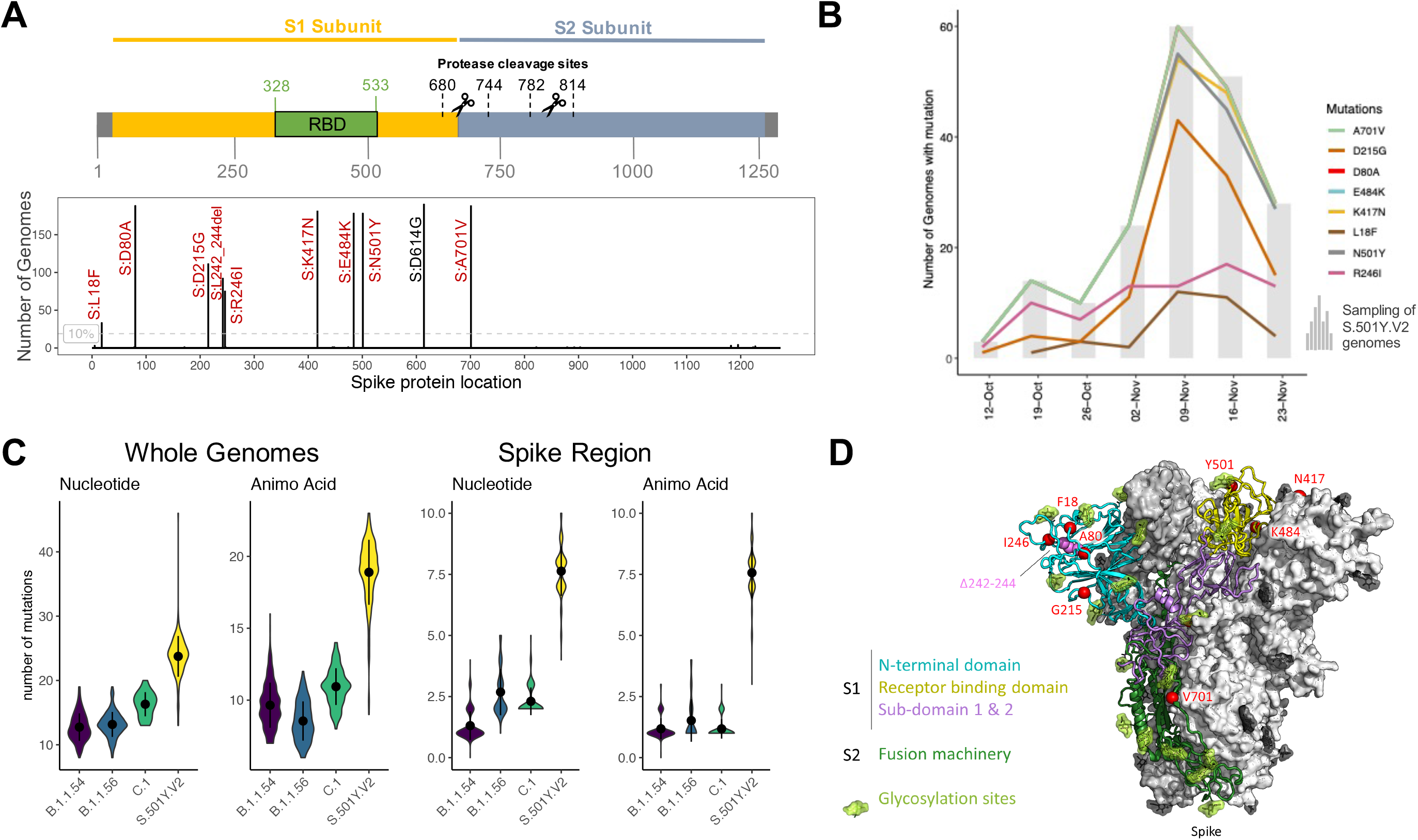
A) Amino acid changes in the spike region of the 190 S501Y.V2 genomes in this study mapped to the spike protein sequences structure, indicating key regions, such as the RBD. Each spike protein variant is shown at their respective protein locations, with the bar lengths representing the number of genomes harboring the specific mutations (Only mutations that appear in >10% of sequences are shown). The D614G mutation (in black) is already present in the parent lineage. B) Changes in the mutation frequency of each variant observed during the course of sampling. Grey bars show the number of S501Y.V2 sequences sampled at a given time point and the colored lines show the change in the number of those sequences harboring each variant at the respective time points. C) Violin plots showing the numbers of nucleotide substitutions and amino acid changes that have accumulated in both the whole genomes and the spike region of the S501Y.V2 lineage, compared to lineages B.1.1.54, B.1.1.56, and C.1, three major lineages circulating in South Africa during the first wave. D) A complete model of the SARS-CoV-2 Spike (S) trimer is shown, with domains of a single protomer shown in cartoon view and coloured cyan (N-terminal domain, NTD), yellow (C-terminal domain/receptor binding domain, CTD/RBD), purple (subdomain 1 and 2, SD1 and SD2), and dark green (S2), while N-acetylglucosamine moieties are coloured light green. The adjacent protomers are shown in surface view and coloured shades of grey. Eight nonsynonymous mutants (red) and a three amino acid deletion (pink) that together define the Spike501Y.V2 lineage are shown with spheres.

### Selection analysis

We examined patterns of nucleotide variations and fluctuations in mutant frequencies at eight polymorphic spike gene sites (Fig. 2B) to determine whether any of the observed polymorphisms might be contributing to changes in viral fitness globally. For this analysis we used 142 037 high quality sequences from GISAID sampled between 24 December 2019 and 14 November 2020, which represented 5964 unique spike haplotypes. The analysis indicated that two of the three sites in RBD (E484 and N501) display a pattern of nucleotide variation that is consistent with the site evolving under diversifying positive selection. The N501Y polymorphism that first appears in our sequences sampled on 15 October shows indication of positive selection on five global tree internal branches, with codon 501 of the spike gene displaying a significant excess of non-synonymous substitutions globally (dN/dS > 1 on internal branches, p=0.0011 by the FEL method), and mutant viruses encoding Y at this site have rapidly increased in frequency in both the UK and in South Africa (Z-score = 11, trend Jonckheere Terpstra non-parametric trend test). Similarly, at codon 484 there is indication of positive selection on seven global tree internal branches, with an overall significant excess of non-synonymous substitutions globally (p=0.015). There is currently no statistical evidence of positive selection at K417. Outside RBD, sites L18 (p<0.001), D80 (p=0.0014), and D215 (p<0.001) show evidence of positive diversifying selection globally with the L18F mutation also having increased in frequency in the regions where it has occurred (Z-score = 17).

## Discussion

We describe and characterise a new SARS-CoV-2 lineage with multiple spike mutations that emerged in a major metropolitan area in South Africa following the first wave of the epidemic and then spread to multiple locations within two other neighbouring provinces. We show that this lineage has rapidly become the dominant circulating genotype, at least in EC and WC, at the time of a rapid resurgence in infections. Whilst the full significance of the mutations is not yet clear, the genomic and epidemiological data suggest increased transmissibility associated with the virus. These data highlight the urgent need to refocus the public health response in South Africa on interrupting transmission, not only to reduce hospitalisations and deaths but to limit the national and international spread of this lineage.

This new lineage has three mutations at key sites in the RBD (K417N, E484K and N501Y). Two of these (E484K and N501Y) are within the receptor-binding motif (RBM), the main functional motif that forms the interface with the human ACE2 (hACE2) receptor. The N501Y mutation has recently been identified in a new lineage in the United Kingdom (B.1.1.7), with some preliminary evidence that it may be more transmissible^29,30^. N501 forms part of the binding loop in the contact region of hACE2, forming a hydrogen bond with Y41 in hACE2^31-33^. It also stabilises K353, one of the virus-binding hotspot residues on hACE2^34^. It is one of the key positions that differentiates SARS-CoV-2 from SARS-CoV and contributes to the enhanced binding affinity of SARS-CoV-2 for hACE2^31,34^. The N501Y mutation has been shown through deep mutation scanning and in a mouse model to enhance binding affinity to hACE2^35,36^. The E484K mutation is uncommon, being present in <0.02% of sequences from outside South Africa. E484 is also in the RBM, and interacts with the K31 interaction hotspot residue of hACE2. There is some evidence that the E484K mutation may modestly enhance binding affinity^35^. K417 is a unique hACE-2 interacting residue that forms a salt bridge interaction across the central contact region with D30 of hACE2^31,32^. This is the most striking difference in the RBD-hACE2 complex between SARS-CoV-2 and SARS-CoV, and contributes to the enhanced binding affinity of SARS-CoV-2 to hACE2^31-33^. Deep mutational scanning suggests that the K417N mutation has minimal impact on binding affinity to hACE2^35^.

The spike RBD is the main target of neutralizing antibodies (NAbs) elicited during SARS-CoV-2 infection^37^. NAbs to the RBD can be broadly divided into four main classes^38^. Of these, class 1 and class 2 antibodies appear to be most frequently elicited during SARS-CoV-2 infection, and their epitopes directly overlap the hACE2 binding site^37^. Class 1 antibodies have a VH3-53 restricted mode of recognition centred around spike residue K417. The K417N mutation would abolish key interactions with class 1 NAbs, and likely contributes toward immune evasion at this site. The N501Y mutation may also have a role in escaping class 1 NAbs, some of which make contact at this site. Class 2 antibodies bind to spike residue E484, and the E484K mutation has been shown to confer resistance to NAbs in this class, and to panels of convalescent sera, suggesting that E484 is a dominant neutralizing epitope^39-42^.

One hypothesis for the emergence of this lineage, given the large number of mutations relative to the background mutation rate of SARS-CoV-2, is that it may have arisen through intra-host evolution in one or more individuals with prolonged viral replication^43,44^. This hypothesis is supported by the long branch length connecting the lineage to the remaining sequences in our phylogenetic tree (Suppl Fig. S10). The N501Y mutation is one of several spike mutations that emerged in an immunocompromised individual in the US who had prolonged viral replication for over 20 weeks^44^. In South Africa, the country with the world’s biggest HIV epidemic, one concern has been the possibility of prolonged viral replication and intra-host evolution in the context of HIV infection, although the limited evidence so far does not suggest that HIV infection is associated with persistent SARS-CoV-2 replication^45^. It should be noted, however, that the observed diversity within this lineage cannot be explained by a single long-term infection in one individual because the lineage contains circulating intermediate mutants with subsets of the main mutations that characterise the lineage. If evolution within long-term infections were the explanation for the evolution of this lineage then one would need to invoke a transmission chain that passes through multiple individuals. Further, antigenic evolution, even within non-immuno-suppressed individuals, could offer an alternative explanation, given that several of the individual sites in spike appear to be under selective pressure worldwide, and that several of the identified mutations have been found in circulating lineages together.

Whilst we have yet to characterise how the mutations (particularly those in the RBM) affect antigenicity, it is plausible that high levels of population immunity could have driven the selection of this lineage. We have very limited SARS-CoV-2 seroprevalence data from South Africa to help understand the true extent of the epidemic. In studies using residual blood samples from routine public sector antenatal and HIV care, seroprevalence in parts of the City of Cape Town was estimated at approximately 40% in July-August, towards the end of the first epidemic wave in that area^46^. We have shown that EC, and Nelson Mandela Bay in particular, were worse affected than City of Cape Town in the first wave, and therefore we believe that population immunity could have been sufficiently high in this region to contribute to population-level selection. Whilst there have been no confirmed re-infections (supported by whole genome sequencing) in South Africa, the true extent of re-infections is unknown and this is now the focus of urgent investigation.

We are now working to understand the phenotypic impact of these SARS-CoV-2 mutations. We are focusing on the following priority studies: we are performing infectivity assays and neutralization assays to understand the effect of these mutations on ACE2 binding and NAb binding; we are analysing clinical data from the three provinces to identify any signals pointing to different disease progression or severity; we have intensified the genomic surveillance in all provinces of South Africa; and we have stepped up monitoring and surveillance for possible re-infections. Whilst the full implications of this new lineage in South Africa are yet to be determined, these findings highlight the importance of coordinated molecular surveillance systems in all parts of the world, to enable early detection and characterisation of new lineages and to inform the global pandemic response.

## Data Availability

All genomic data has been deposited in GISAID

## Acknowledgements

This research reported in this publication was supported by the Strategic Health Innovation Partnerships Unit of the South African Medical Research Council, with funds received from the South African Department of Science and Innovation.

## Author Contributions

Produced SARS-CoV-2 genomic data: Jennifer Giandhari, Sureshnee Pillay, Susan Engelbrecht and Arshad Ismail

Collected samples and curated metadata: Nokukhanya Msomi, Koleka Mlisana, Nei-yuan Hsiao, Denis York, Dominique Goedhals, Anne von Gottberg, Sibongile Walaza, Allison J. Glass, Inbal Gazy, Alex Sigal, Gert Van Zyl, Wolfgang Preiser, Diana Hardie

Analysed the data: Houriiyah Tegally, Eduan Wilkinson, Marta Giovanetti, Richard J Lessells, Sergei L Kosakovsky Pond, Steven Weaver, Darren Martin, ennifer Giandhari, Sureshnee Pillay, Emmanuel James San, Susan Engelbrecht, Francesco Pettruccione, Arshad Ismail, Jinal N. Bhiman, Vagner Fonseca, José Lourenço, Luiz Carlos Junior Alcantara, and Tulio de Oliveira

Helped with data interpretation: Diana Hardie, Nei-yuan Hsiao, Darren Martin, Dominique Goedhals, Emmanuel James San1, Marta Giovanett, José Lourenço, Luiz Carlos Junior Alcantara, Tulio de Oliveira

Initial Manuscript Writing: Richard J Lessells, Houriiyah Tegally, Eduan Wilkinson, Marta Giovanetti, Darren Martin and Tulio de Oliveira

Review of the manuscript: All authors.

