## Supplementary Material for "Emergence and rapid spread of a new severe acute respiratory syndrome-related coronavirus 2 (SARS-CoV-2) lineage with multiple spike mutations in South Africa"

**Supplementary Figures: Fig S1-S11**

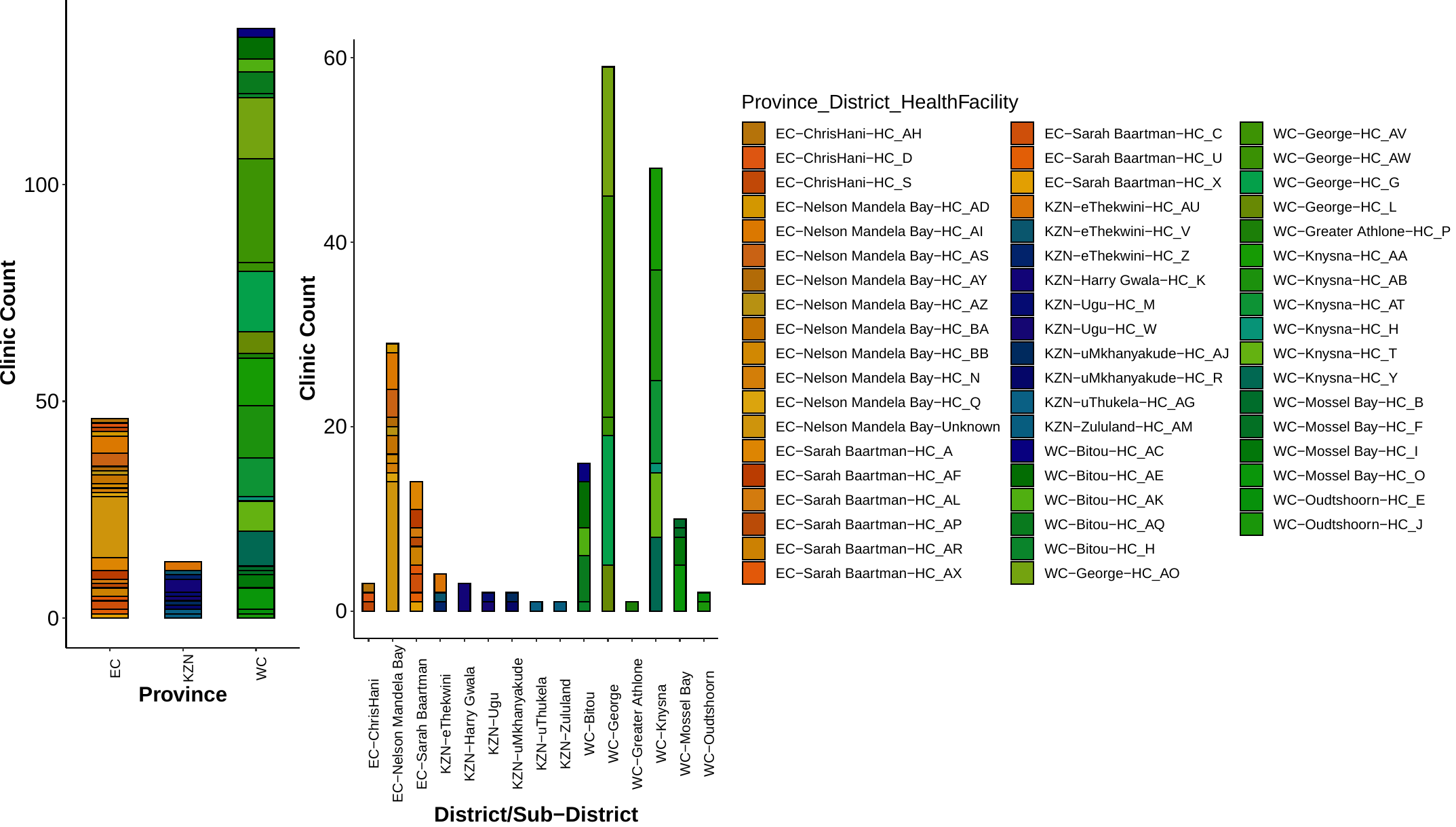

**Supplementary Fig S1:** A representation of the number of clinics per province where the S501Y.V2 lineage was detected in sampled genomes.

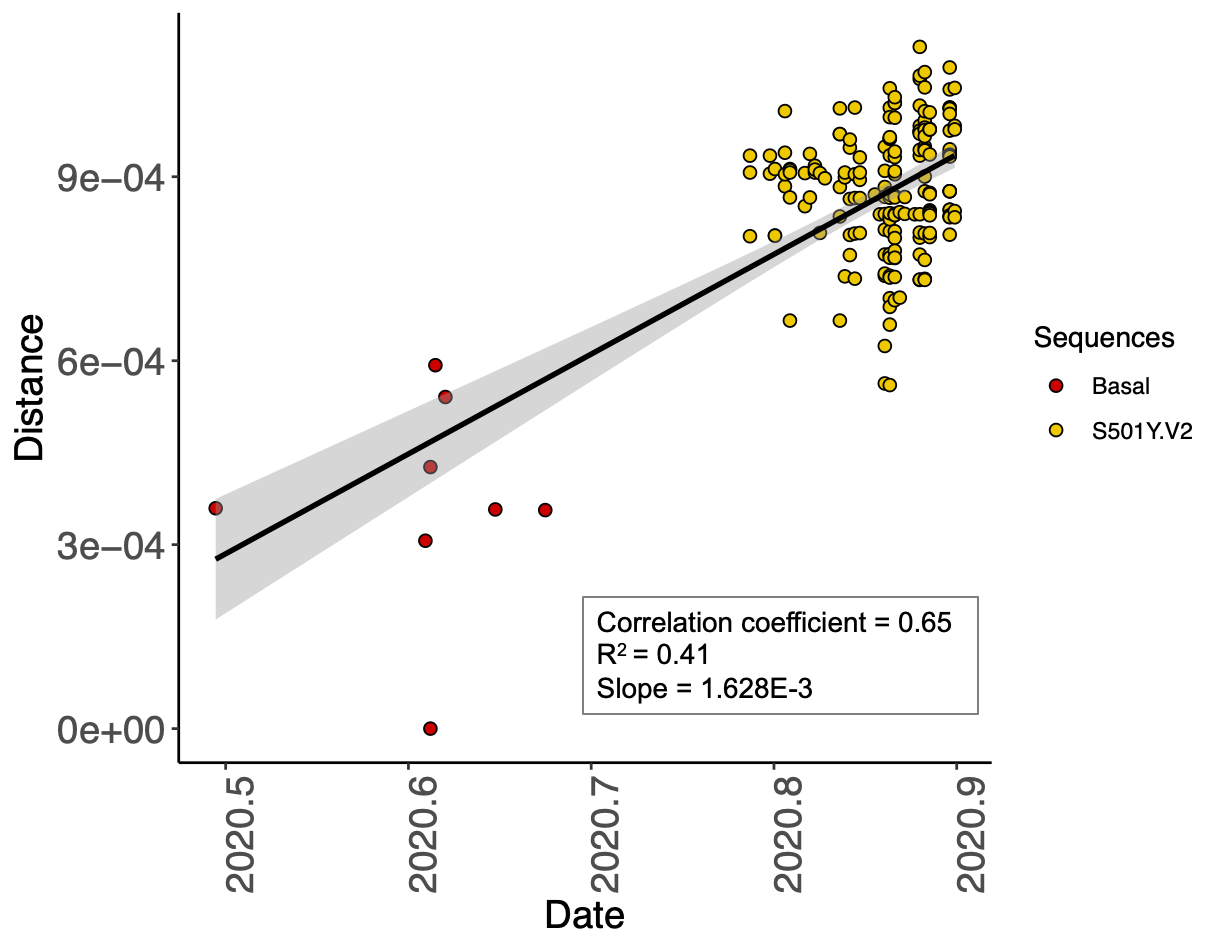

**Supplementary Fig S2:** Temporal signal obtained from TempEst analysis for the S501Y.V2 lineage cluster (yellow), including its basal sequences (red), showing a relatively strong clock-like behavior (correlation coefficient = 0.65, R^2^ = 0.41) and a regression line slope, representing mean evolutionary rate, of 1.628E-3 nucleotide changes/site/year.

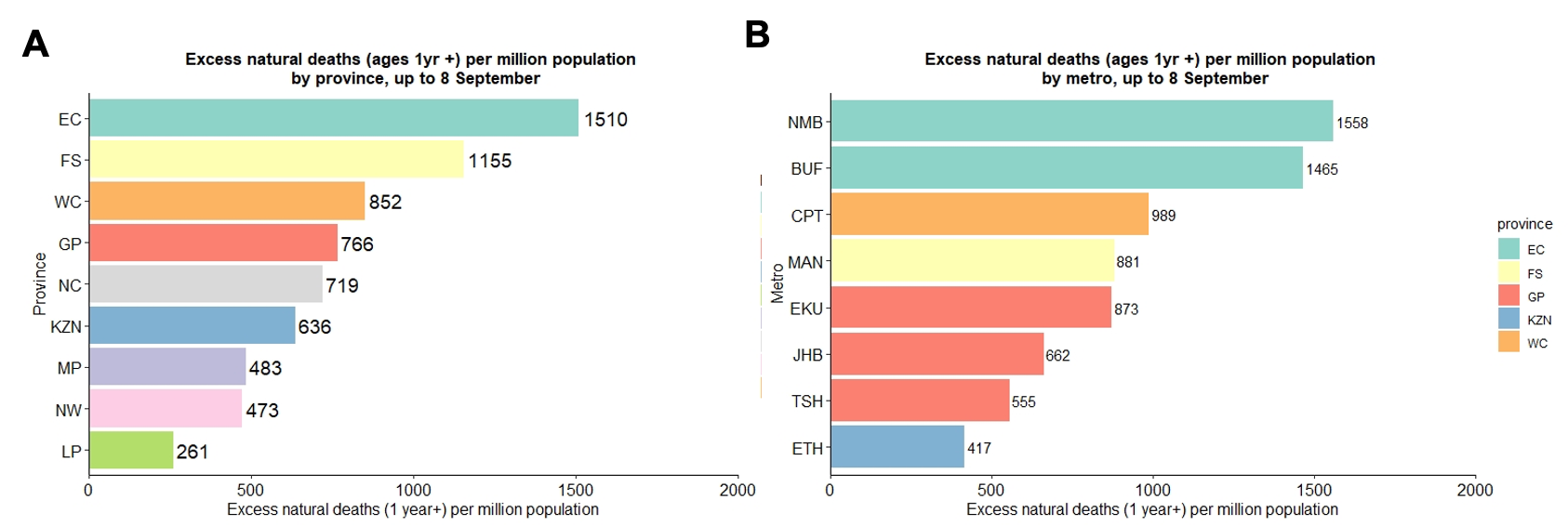

**Supplementary Fig S3:**  Excess deaths per million population, by province and metropolitan municipalities of South Africa, up until the week ending 8 September 2020 (just past the first epidemic peak). These graphs indicate the disproportionate impact of the first wave of the epidemic in the province of the Eastern Cape (A) and its metro areas Nelson Mandela Bay (NMB) and Buffalo City (BUF) (B). [Key: EC=Eastern Cape, FS=Free State, WC=Western Cape, GP=Gauteng, NC=Northern Cape, KZN=KwaZulu-Natal, MP=Mpumalanga, NW=North West; NMB=Nelson Mandela Bay, BUF=Buffalo City, CPT=Cape Town, MAN=Mangaung, EKU=Ekurhuleni, JHB=Johannesburg, TSH=Tshwane, ETH=Ethekwini]

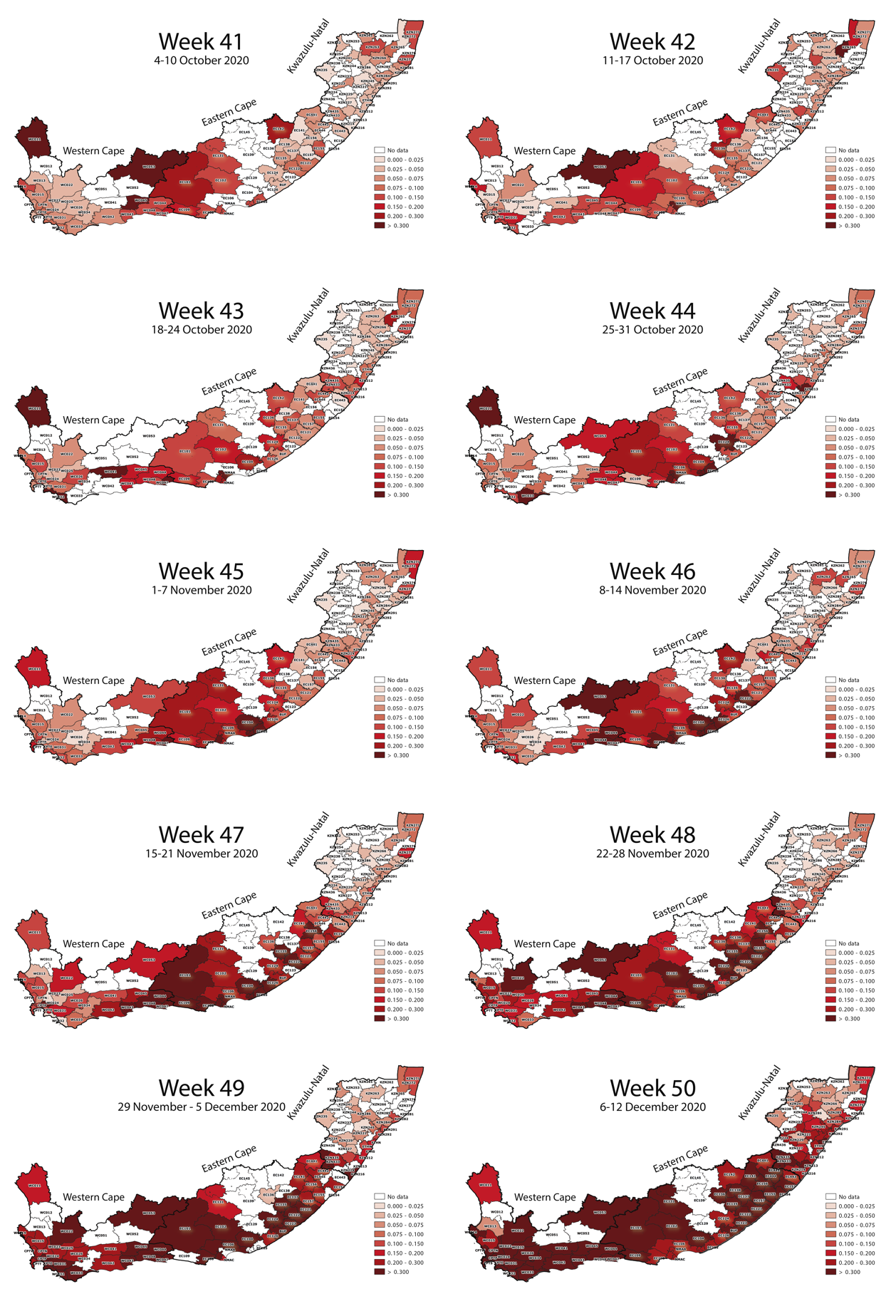

**Supplementary Fig S4:** Geographical maps of the Western Cape, the Eastern Cape and KwaZulu-Natal, the three main provinces investigated in this study), showing a weekly progression of SARS-CoV-2 prevalence per district and colored by the rate of positive SARS-CoV-2 PCR-tests per district. Data for this figure was obtained from the NICD weekly testing report.

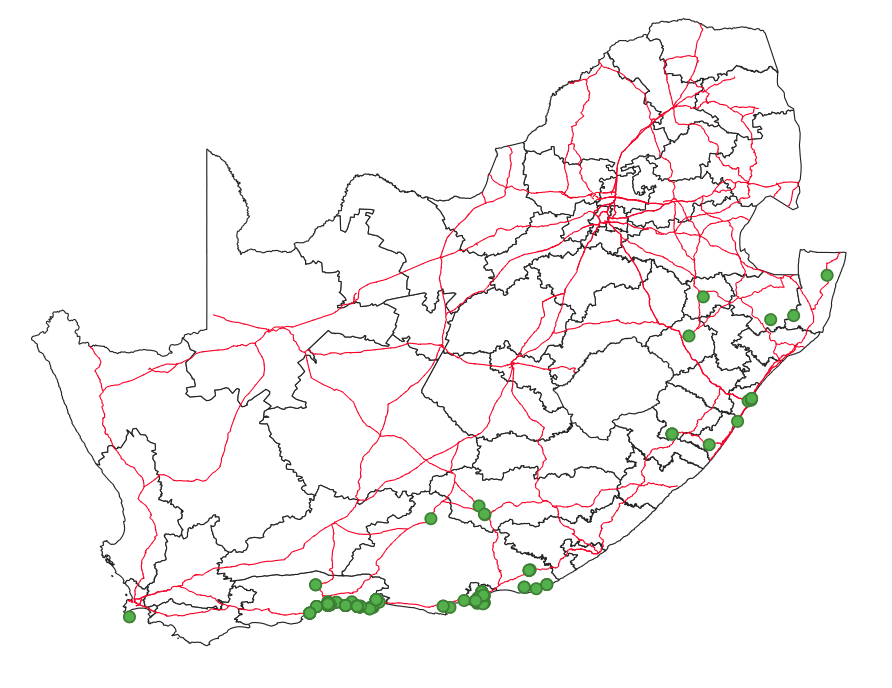

**Supplementary Fig S5:** A general map of South Africa, showing the sampling location of the S501Y.V2

genomes in this study (green dots) in relation to the main road networks in the country, hinting to potential land transmission routes of this lineage along the coast.

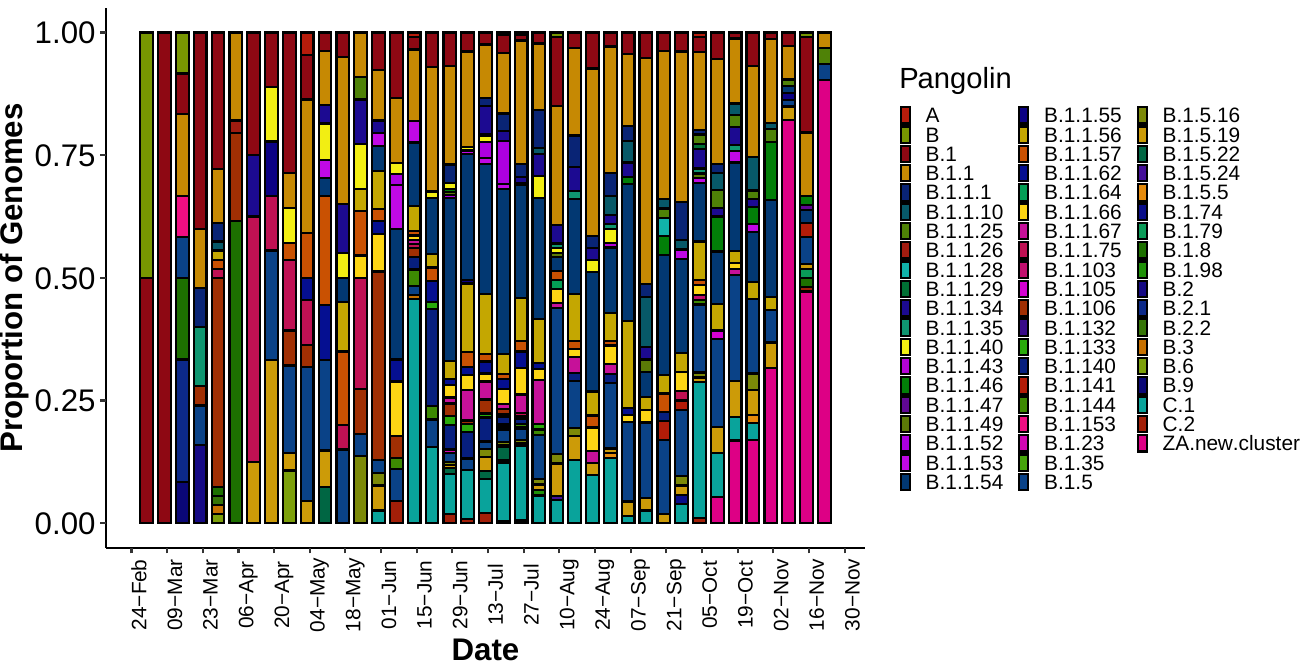

**Supplementary Fig S6:** Progression of SARS-CoV-2 Pangolin lineages circulating in South Africa from March to December, showing over-representation of the new S501Y.V2 lineage from October onwards (in pink)

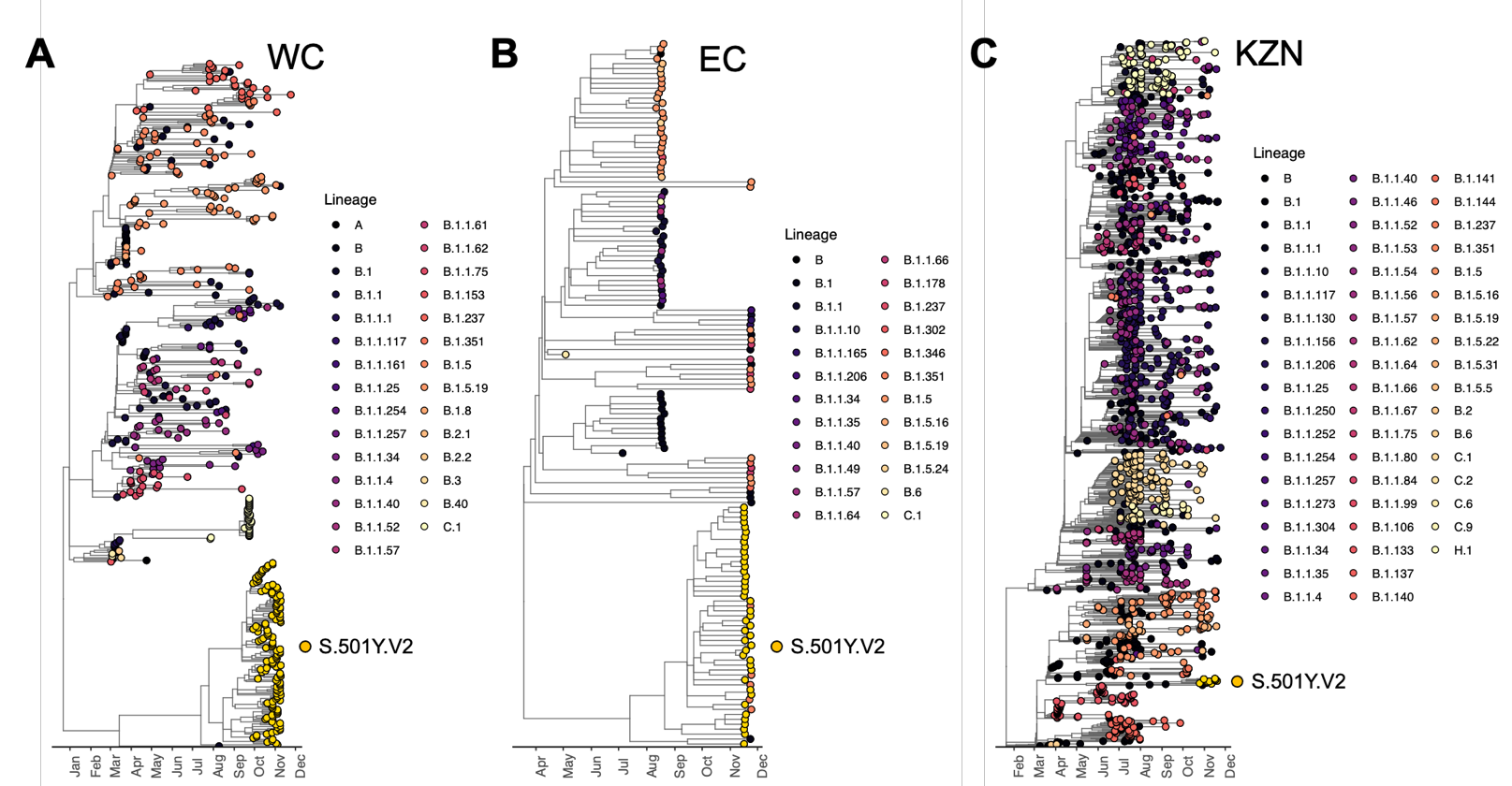

**Supplementary Fig S7:** Independent regional phylogenetic trees for WC, EC and KZN, showing a variety of circulating lineages prior to October and the dominance of S.501Y.V2 (in yellow) in late October and November, especially in WC and EC.

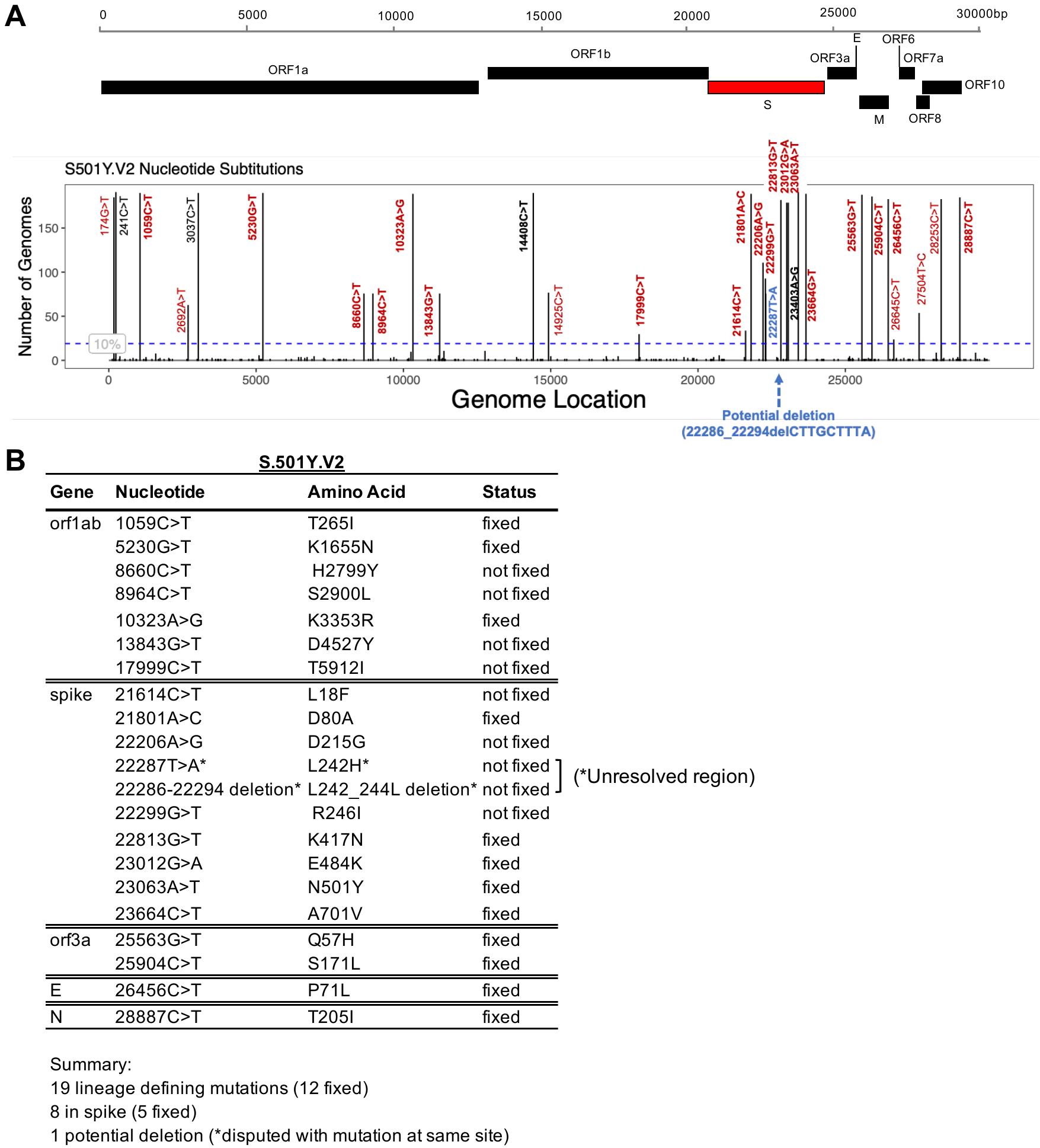

**Supplementary Fig S8:** Overview of mutations associated with the S501Y.V2 lineage. A) All nucleotide substitutions present in >10% of the genomes in the S501Y.V2 lineage mapped on the SARS-CoV-2 genomic structure. Mutations present in the parent lineage (B.1) are marked in black, while mutations specific to this lineage are marked in red or blue. All non-synonymous mutations are styled in bold, and also reported in panel B. The site in blue is unresolved for the moment as we detect a non-synonymous mutation in some sequences/reads and a 9-nucleotide long deletion in others. B) A summary of all non-synonymous lineage-defining changes in relevant genes occurring in the S501Y.V2 lineage. The unresolved site is marked with an asterisk (*).

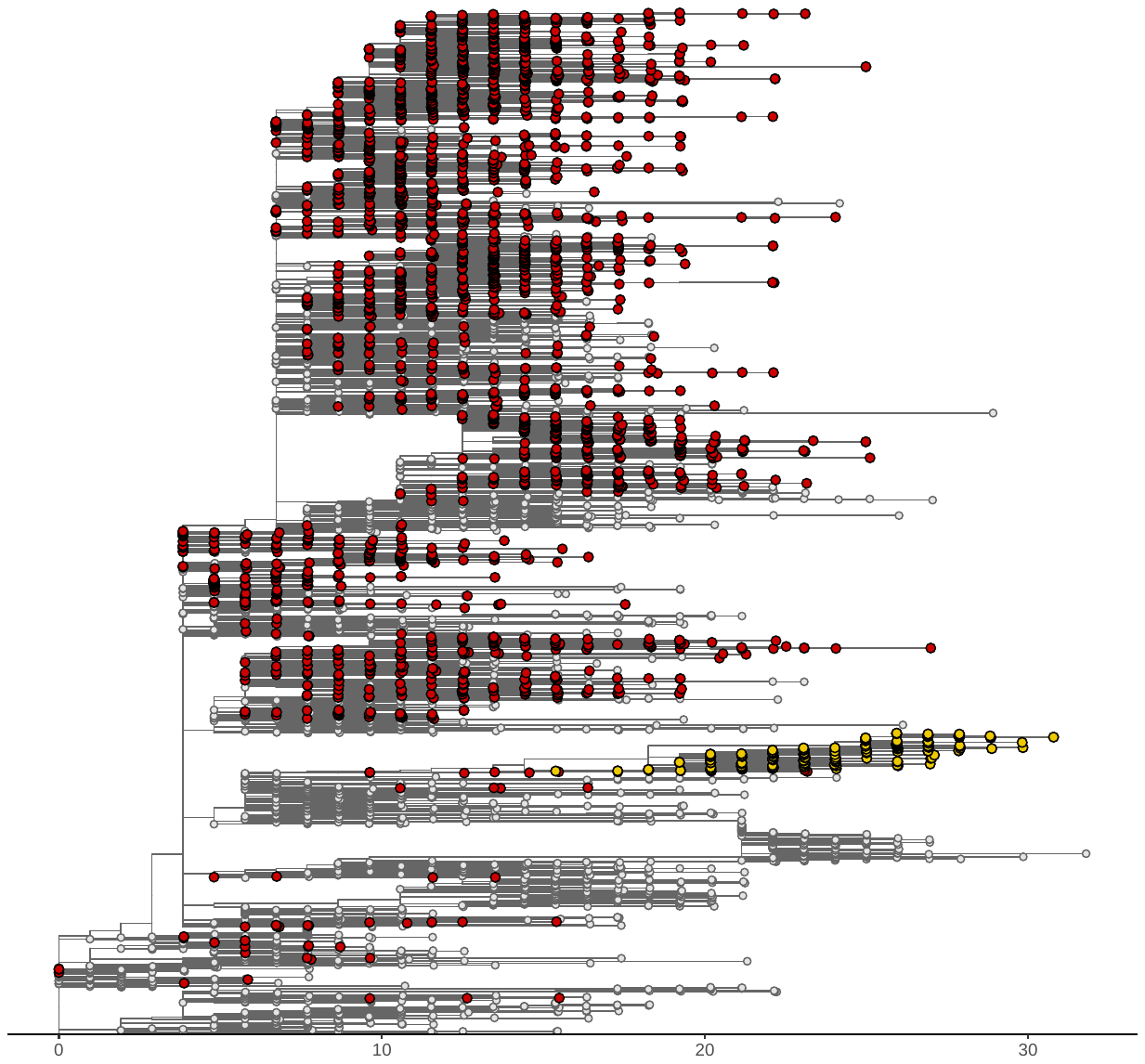

**Supplementary Figure S9:** A Maximum-Likelihood tree of 2285 South African genomes and an equal number of global genomes, where branch lengths represent the diversity of the genomes against the Wuhan reference. The S.501Y.V2 lineages (in yellow) shows relatively longer branches compared to other South Africa viral genomes, forming other lineages circulating in the country, prior to the detection of this new lineage

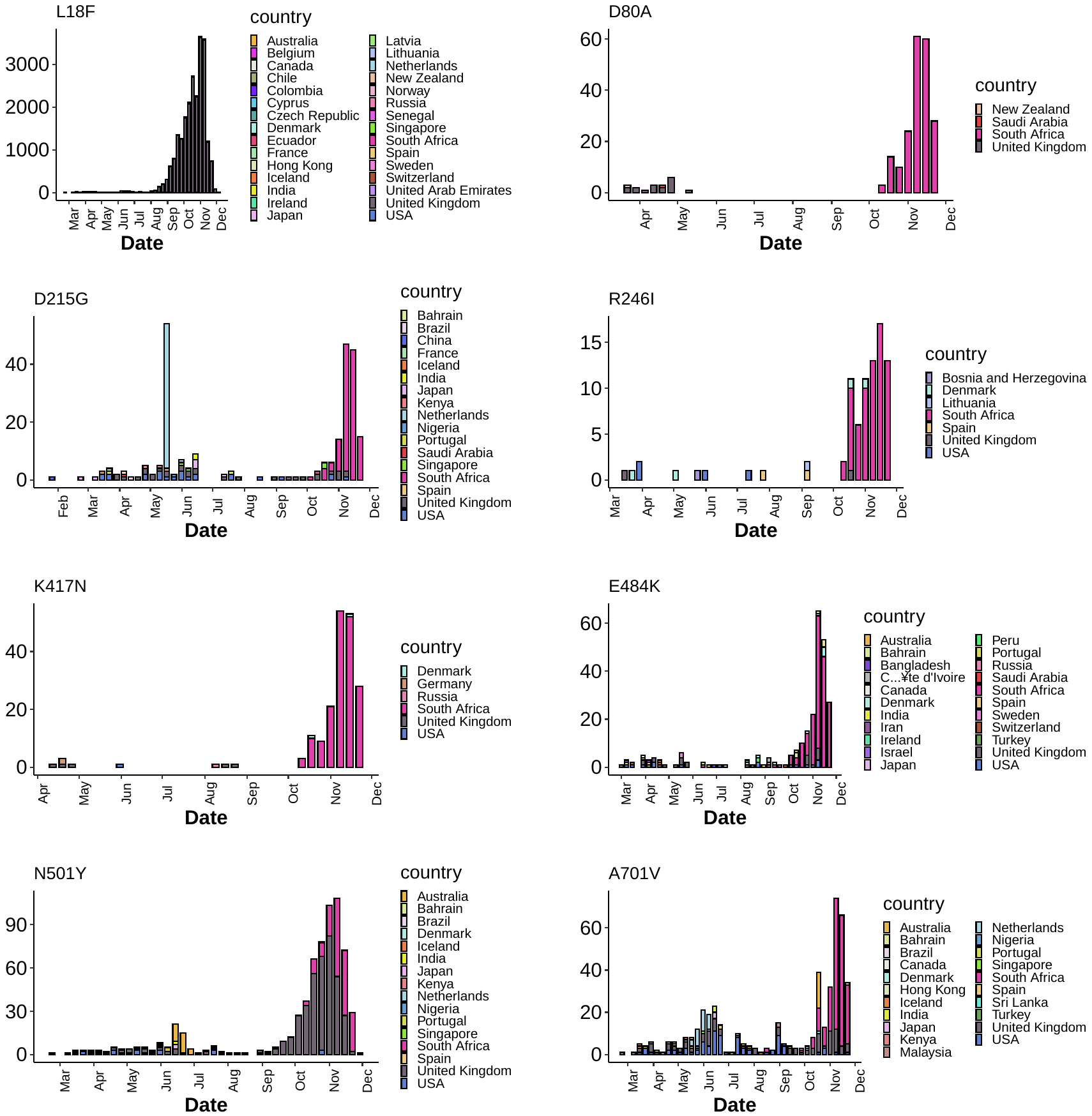

**Supplementary Fig S10:** Showing the prevalence of the 8 spike mutations around the world, indicating that several of them emerge independently in multiple regions.

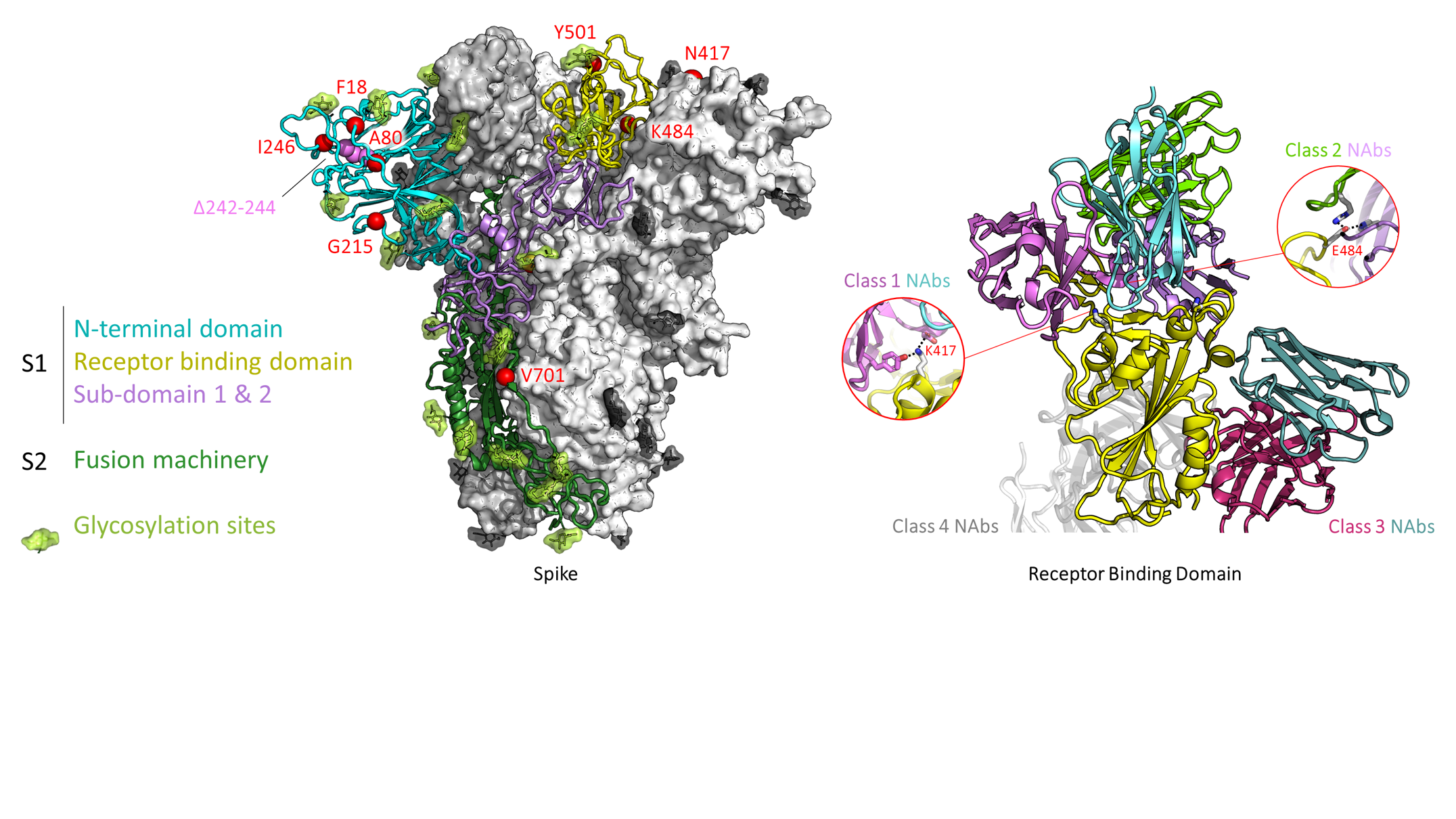

**Supplementary Fig S11:** A model of the SARS-CoV-2 receptor binding domain in cartoon view (yellow), showing representative Fab domains for neutralising antibodies from classes 1, 2, 3, and 4. Two zoomed insets show common, key interactions between RBD residue K417 and class 1 NAbs, or RBD residue E484 and class 2 NAbs.
